# A Systematic Review of ChatGPT and Other Conversational Large Language Models in Healthcare

**DOI:** 10.1101/2024.04.26.24306390

**Authors:** Leyao Wang, Zhiyu Wan, Congning Ni, Qingyuan Song, Yang Li, Ellen Wright Clayton, Bradley A. Malin, Zhijun Yin

**Affiliations:** Department of Computer Science, Vanderbilt University, Nashville, TN, USA, 37212; Department of Biomedical Informatics, Vanderbilt University Medical Center, TN, USA, 37203; Department of Pediatrics, Vanderbilt University Medical Center, Nashville, Tennessee, USA, 37203; Center for Biomedical Ethics and Society, Vanderbilt University Medical Center, Nashville, Tennessee, USA, 37203; Department of Biostatistics, Vanderbilt University Medical Center, TN, USA, 37203

**Keywords:** large language model, ChatGPT, artificial intelligence, natural language processing, healthcare, summarization, medical knowledge inquiry, reliability, bias, privacy

## Abstract

**Background:** The launch of the Chat Generative Pre-trained Transformer (ChatGPT) in November 2022 has attracted public attention and academic interest to large language models (LLMs), facilitating the emergence of many other innovative LLMs. These LLMs have been applied in various fields, including healthcare. Numerous studies have since been conducted regarding how to employ state-of-the-art LLMs in health-related scenarios to assist patients, doctors, and public health administrators.

**Objective:** This review aims to summarize the applications and concerns of applying conversational LLMs in healthcare and provide an agenda for future research on LLMs in healthcare.

**Methods:** We utilized PubMed, ACM, and IEEE digital libraries as primary sources for this review. We followed the guidance of Preferred Reporting Items for Systematic Reviews and Meta-Analyses (PRIMSA) to screen and select peer-reviewed research articles that (1) were related to both healthcare applications and conversational LLMs and (2) were published before September 1^st^, 2023, the date when we started paper collection and screening. We investigated these papers and classified them according to their applications and concerns.

**Results:** Our search initially identified 820 papers according to targeted keywords, out of which 65 papers met our criteria and were included in the review. The most popular conversational LLM was ChatGPT from OpenAI (60), followed by Bard from Google (1), Large Language Model Meta AI (LLaMA) from Meta (1), and other LLMs (5). These papers were classified into four categories in terms of their applications: 1) summarization, 2) medical knowledge inquiry, 3) prediction, and 4) administration, and four categories of concerns: 1) reliability, 2) bias, 3) privacy, and 4) public acceptability. There are 49 (75%) research papers using LLMs for summarization and/or medical knowledge inquiry, and 58 (89%) research papers expressing concerns about reliability and/or bias. We found that conversational LLMs exhibit promising results in summarization and providing medical knowledge to patients with a relatively high accuracy. However, conversational LLMs like ChatGPT are not able to provide reliable answers to complex health-related tasks that require specialized domain expertise. Additionally, no experiments in our reviewed papers have been conducted to thoughtfully examine how conversational LLMs lead to bias or privacy issues in healthcare research.

**Conclusions:** Future studies should focus on improving the reliability of LLM applications in complex health-related tasks, as well as investigating the mechanisms of how LLM applications brought bias and privacy issues. Considering the vast accessibility of LLMs, legal, social, and technical efforts are all needed to address concerns about LLMs to promote, improve, and regularize the application of LLMs in healthcare.

## Introduction

Since Chat Generative Pre-trained Transformer (ChatGPT) was released on November 30^th^, 2022, extensive attention has been drawn to generative AI and large language models (LLMs) [1]. ChatGPT is a representative conversational LLM that generates text based on its training on an extremely large amount of data from mostly the public domain [1]. Modern LLMs (such as GPT-4) incorporate in-text learning, which enables them to interpret and generalize user inputs in the form of natural language prompts that require little to no fine-tuning [2]. These LLMs have surpassed the limits of prior incarnations and are now capable of performing various complex natural language processing (NLP) tasks, including translation and question-answering [3]. In comparison to traditional chatbots, the current array of conversational LLMs can generate seemingly human-like coherent texts [3]. Moreover, since these models are trained on publications from online libraries such as Common Crawl and Wikipedia, they can generate seemingly scientific and competent answers [4].

Due to the high quality of their responses and the broad training database of modern LLMs, a growing body of studies has emerged regarding the applications of chatbots, particularly ChatGPT, in the domain of health and medicine [5]. However, most LLMs are not specially designed for healthcare and, as a result, certain practical pitfalls may exist when they are put into practice in that setting. Thus, there is a need to compile the latest achievements in this domain so that potential issues and guidance for new research directions can be laid out. Several reviews have been published to discuss the appropriateness of a particular application of LLMs in a specific aspect [1,6–10] but none of them summarized the overall problems systematically [8]. For example, Huang et al. summarized only the application of ChatGPT in dentistry without considering the broader landscape of other subfields in healthcare [6]. Mesko and Topol only discussed regulatory challenges, neglecting concerns about reliability in their application, such as their accuracy and consistency of responses [10]. Wang et al. discussed the ethical considerations of using ChatGPT in healthcare [7], they did not consider other LLMs for analysis, account for other common challenges such as reliability, or mention detailed applications of the models. While Snoswell et al. reviewed applications of LLMs in medicine, they did not conduct a systematic review [9]. Moreover, their work focused on LLMs’ educational and research applications rather than their clinical usage. Although Sallam conducted a systematic review [8], the articles Sallam considered were mostly editorials, letters to the editors, opinions, commentaries, news articles, and preprints, as opposed to research articles. In addition, Sallam focused on educational and research applications of ChatGPT only.

This review focuses on peer-reviewed research articles on conversational LLMs that emerged after ChatGPT, which was initially based on GPT-3, and their applications in healthcare. We aim to summarize the applications of conversational LLMs in the field of healthcare with concrete experiments and identify potential concerns about using such LLMs in this field that need to be addressed in the future.

## Methods

We searched for papers that contained at least one word associated with LLMs {“ChatGPT”, “LLaMA”, “GPT-3”, “LaMDA”, “PalM”, “MT-NLG”, “GATO”, “BLOOM”, “Alpaca”, “Large Language Model”} and at least one word associated with healthcare {“health”, “diagnosis”, “intervention”, “patient”} published before September 1^st^, 2023 on PubMed, Association for Computing Machinery (ACM) Digital Library, and Institute of Electrical and Electronics Engineers (IEEE) Xplore. This systematic review applied the Preferred Reporting Items for Systematic Reviews and Meta-Analyses (PRIMSA) guidelines to steer the paper search [11]. Relevant publications were gathered and downloaded on September 3^rd^, 2023. For simplicity, all the LLMs mentioned henceforth refer to conversational LLMs.

The inclusion criteria for a paper are that 1) it was published as a peer-reviewed scientific research article between November 1^st^, 2022, and September 1^st^, 2023, and 2) it focuses on applications of LLMs in addressing a healthcare-related problem, which includes, but is not limited to, promotion of personal or public health and well-being or the potential to alleviate the workload of healthcare providers. We excluded a paper if it was (1) not a peer-reviewed research article; (2) not related to healthcare applications (e.g., LLMs applied to preparing manuscripts for peer-review); (3) not accessible; (4) a duplicate of another paper considered; or (5) about LLMs released before GPT-3, such as BERT. We excluded BERT-related papers because this LLM, which was built upon the encoder of a transformer, is mainly applied in fine-tuning downstream machine-learning tasks. While the implementation of a chatbot based on BERT is feasible, it waned in popularity as an LLM after the introduction of ChatGPT, which was built upon the decoder of a transformer. The complete set of papers meeting the criteria were downloaded from the three digital libraries for further screening. Specifically, five of the authors of this review (LW, ZW, CN, QS, and YL) participated in paper screening and summarization under the supervision of the corresponding author ZY. A screening protocol was created collectively after the team jointly reviewed 50 randomly selected papers. Each unreviewed paper was then screened by not fewer than two authors based on the protocol. All the papers in the final collection were summarized by the co-authors according to their LLM applications in healthcare and the concerns raised.

## Results

Figure 1 demonstrates the paper selection process. The initial keyword search identified a total of 820 articles, with 736 articles from PubMed, 49 papers from ACM Digital Library, and 35 papers from IEEE Xplore. The evaluation of the 820 articles was distributed among the authors for screening the titles and abstracts. The inter-rater reliability was assessed by computing a Kappa score, yielding a value of 0.72. After screening, we excluded 599 articles from PubMed, 46 articles from ACM Digital Library, and 33 papers from IEEE Xplore because they were either not relevant to the research topic or not research articles. Next, we extracted the full papers of the remaining 142 research articles and manually examined them for the five excluding criteria (See Methods). This led to a final set of 65 papers for full-paper review and summarization - 63, 2, and 0 from PubMed, ACM Digital Library, and IEEE Xplore, respectively. Among these selected papers, 60 were related to ChatGPT from OpenAI, 1 was related to Large Language Model Meta AI (LLaMA) from Meta, 1 was related to Bard based on Language Model for Dialogue Applications (LaMDA) from Google, and 5 of them are related to other LLMs (See Supplemental Table 1).

**Figure 1.**
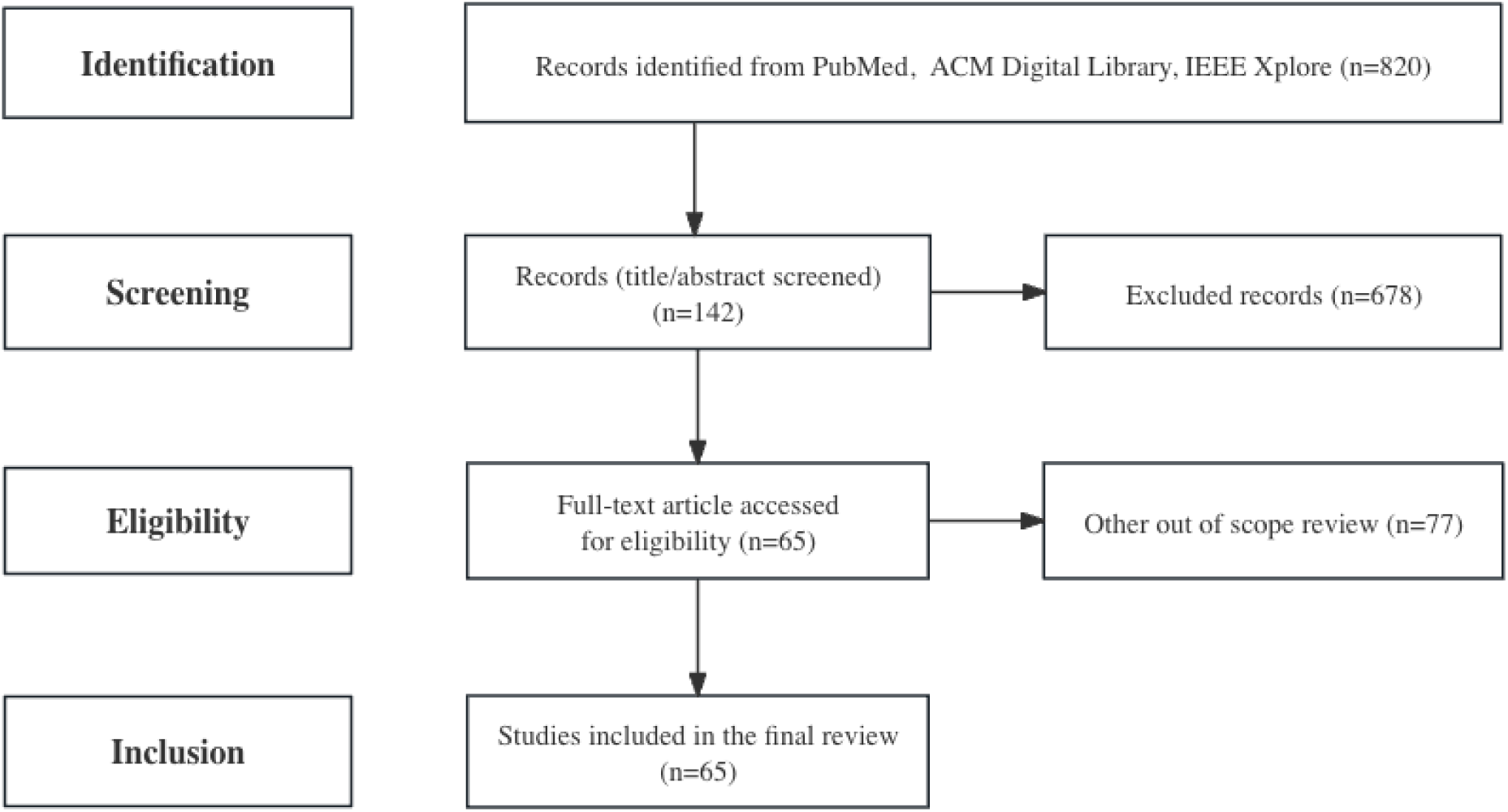
A flowchart of the article selection process based on the PRIMSA guidelines. PRIMSA: Preferred Reporting Items for Systematic Reviews and Meta-Analyses; ACM: Association for Computing Machinery; IEEE: Institute of Electrical and Electronics Engineers.

Figure 2 illustrates the main topics of applications and concerns mentioned by the reviewed papers on applying LLMs in healthcare settings. The multifaceted applications of LLMs can be divided into four primary categories: Summarization, Medical Knowledge Inquiry, Prediction, and Administration.

- Summarization (25 papers): LLMs are potential tools for summarizing complex information or documentation in clinical domains.
- Medical Knowledge Inquiry (30 papers): LLMs demonstrate proficiency in answering a diverse array of medical questions and/or examinations, which enhance public access to medical knowledge.
- Prediction (22 papers): LLMs demonstrate high *diagnostic* accuracy in multiple medical scenarios (15 papers), offer virtual assistance in diverse *treatments* (12 papers), and excel in predicting drug interactions and *synergies* (1 paper).
- Administration (9 papers): LLMs streamline various tasks, including *documentation* (5 papers) and *information collection* (5 papers) to monitor the trend of public health.

**Figure 2.**
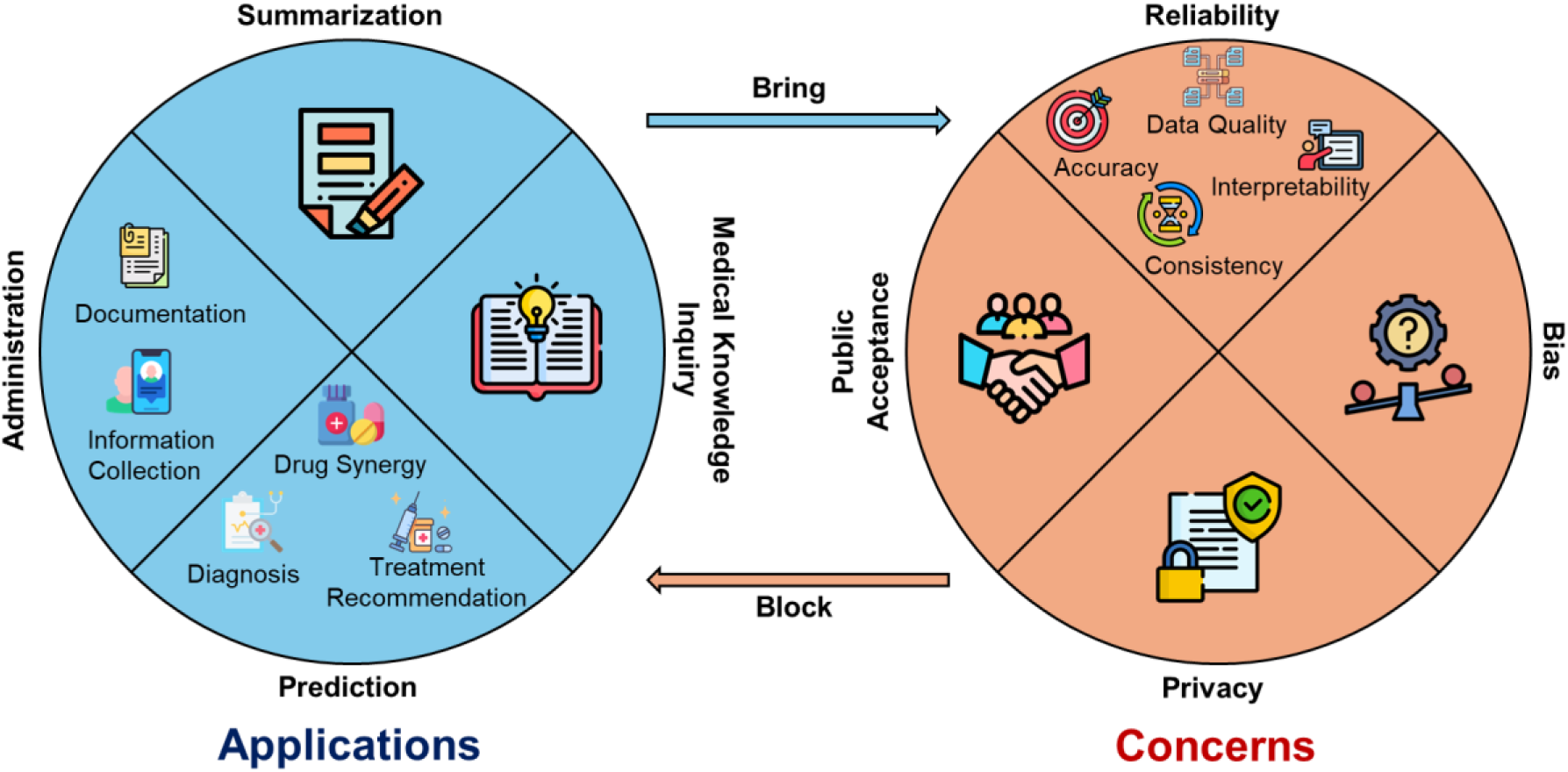
A summary of the applications and concerns about LLMs in healthcare as communicated by the reviewed papers. LLMs: large language models.

The concerns surrounding the application of LLMs in healthcare were varied, each with nuanced considerations.

- Reliability (55 papers): This includes *accuracy* (45 papers), or the correctness of the responses from LLMs; *consistency* (13 papers), whether LLMs produce the same response to the same questions with different prompts; *interpretability* (5 papers), whether LLMs can explain their responses well and the *data quality* of the training dataset (16 papers).
- Bias (16 papers): The applications of LLMs may result in biased responses, which will exacerbate disparity and inequality in healthcare, particularly in terms of *financial costs* (1 paper), *readability* (5 papers), and *accessibility* (3 papers).
- Privacy (6 papers): Training LLMs in healthcare settings requires a large number of health data which, however, is sensitive and may bring privacy issues.
- Public Acceptance (4 papers): Building trust in LLMs from the public is pivotal for widespread acceptance and usage of LLM-based healthcare applications.

### Applications

All reviewed research papers demonstrated the usability or tested the capability of LLMs for healthcare applications in clinical or research domains, which can be further classified into the following four categories: summarization, medical knowledge inquiry, prediction, and administration.

#### Summarization

ChatGPT has been shown to be effective in summarizing medical documents for a diverse set of applications [13,14], including tasks such as adapting clinical guidelines for diagnosis, treatment, and disease management [15], summarizing medical notes [16–18], assisting in writing medical case reports [19–24], and generating and translating radiological reports [18,25]. Notably, efforts have been made to integrate ChatGPT-4^1^ with “Link Reader” for automating medical text synthesis [26], which boosted model performance in providing answers according to clinical guidelines [26]. Another study [23] explored ChatGPT’s role in supporting healthcare professionals in creating medical reports from real patient laboratory results to offer treatment recommendations based on patients’ health conditions [23].

ChatGPT proved beneficial for summarizing research papers as well [27]. Notably, it demonstrated impressive performance in summarizing conference panels and recommendations [27], generating research questions [28], extracting data from literature abstracts [29], drafting medical papers based on given datasets [30], and generating references from medical articles [31]. ChatGPT was also utilized to evaluate the quality and readability of online medical text regarding shockwave therapy for erectile dysfunction [32]. These applications highlighted the potential of LLMs to condense complex and extensive research materials, allowing for more accessible comprehension and utilization of information in healthcare.

#### Medical Knowledge Inquiry

ChatGPT can be applied to answer questions about healthcare, as evidenced by its excellent performance in various studies [26,33–42]. For instance, ChatGPT has shown remarkable accuracy in reasoning questions and medical exams [43,44], even successfully passing the Chinese Medical Licensing Exam [45] and the United States Medical Licensing Exam (USMLE) [46]. It also performed well in addressing radiation oncology physics exam questions [47]. Likewise, “ChatGPT would have been at the 87^th^ percentile of Bunting’s 2013 international cohort for the Cardiff Fertility Knowledge Scale and at the 95^th^ percentile on the basis of Kudesia’s 2017 cohort for the Fertility and Infertility Treatment Knowledge Score” [48]. In addition, ChatGPT showed promising results in a simulated Ophthalmic Knowledge Assessment Program (OKAP) exam [49]. However, the average score of ChatGPT was 60.17% in the Membership of the Royal College of General Practitioners Applied Knowledge Test (AKT), which is below 70.4%, the mean passing threshold in the last 2 years [50].

Furthermore, LLMs have been shown to be effective at making medical knowledge accessible to the public. In particular, a fine-tuned chatbot based on LLaMA demonstrated enhanced performance in identifying patients’ needs and providing informed suggestions [51]. In the realm of medical advice, ChatGPT generated educational documents, answered questions about allergy and immunology [52], and countered vaccine conspiracy theories [53]. It can also answer the most frequently asked questions about the COVID-19 pandemic. Its overall responses to queries related to cognitive decline were equivalent to and, at times, more reliable than Google’s [54]. According to Bulck and Moons [55], in comparison to Google search, 40% of the 20 experts (19 nurses; 1 dietitian) considered answers from ChatGPT of greater value, 45% regarded them as equal value and 15% deemed them less valuable. Therefore, many experts predicted that patients will gradually rely more on LLMs (particularly ChatGPT) and less on Google searches due to the high quality and accessibility of the answers from LLMs. Regarding cancer myths and misconceptions, 97% of expert reviews deemed answers from ChatGPT to be accurate [56]. In addition, Bird and Lotfi optimized a chatbot that could answer mental health-related questions with an accuracy of 89% [57]. Overall, LLMs, particularly ChatGPT, demonstrate an impressive performance in public education in health.

#### Prediction

LLMs have been shown to have predictive capabilities in diagnosis, treatment recommendations, and drug interactions and synergies.

### Diagnosis

ChatGPT has exhibited the potential to achieve high accuracy in diagnosing specific diseases [14,58], providing diagnostic suggestions in simulated situations [35,59] or using given lab reports for diagnosis [60]. ChatGPT has been evaluated in dental [6], allergy [52], and mental disorders diagnoses [61]. Particularly, GPT-3 can be used to differentiate Alzheimer’s patients from healthy controls using speech data [61]. Beyond ChatGPT, other generative AI frameworks, such as DR.BENCH [62], were employed for clinical diagnostic reasoning tasks [62]. Moreover, various pre-trained LLMs can extract microbe-disease relationships from biomedical texts in zero-shot/few-shot contexts with high accuracy, with an average F1 score, precision, and recall greater than 80% [63]. In addition, ChatGPT was the best LLM when predicting high acuity cases than predicting low acuity cases according to emergency severity index (ESI), with a sensitivity of 76.2%, a specificity of 93.1%, compared to the overall sensitivity of 57.1%, specificity of 34.5% [64].

For example, Hirosawa and colleagues [4] obtained ChatGPT’s diagnostic response by describing a clinical scenario. The prompt began with “Tell me the top 10 suspected illnesses for the following symptoms”; Then, patients’ personal information (e.g., age and family history) was provided in this prompt along with other clinical data (e.g., symptoms, medication, and physical examination). According to the study, the top ten suspected diseases generated by ChatGPT achieved a rate of 93% (28/30) in overall correctness. While such a level of performance is impressive, physicians still made a better prediction than ChatGPT. With respect to the top five diagnoses, physicians achieved an accuracy of 98% while ChatGPT only achieved 83%. As to the top suspected disease, ChatGPT only had a correct rate of 53.3%, versus 98.3% achieved by physicians [4].

### Treatment recommendations

LLMs can offer treatment recommendations while listing the side effects of these treatments [58]. They have been involved in the treatment of various diseases such as allergy and immunology [52]. ChatGPT can identify guideline-based treatments for advanced solid tumors [65], such as breast tumor treatment [66]. LLMs can also assist with treatment planning [67], and brain glioma adjuvant therapy decision-making [21]. Similarly, NYUTron, a large language model trained on unstructured clinical notes, has been applied for clinical predictive tasks in treatments [19]. ChatGPT can effectively recommend breast tumor management strategies based on clinical information from ten patients [66], enhance clinical workflow, and assist in responsible decision-making in pediatrics [12]. In addition, ChatGPT can recommend cancer screening given the radiology reports, with an accuracy of 88% [68]. Overall, ChatGPT performs well in certain scenarios of disease prevention and screening recommendations.

### Drug synergies

LLMs also demonstrate high utility when characterizing drug effects. Notably, ChatGPT was employed to predict and explain drug-drug interactions [69]. In this study, the LLMs were asked about pairing or interaction between drugs, and their responses are evaluated in terms of correctness and conclusiveness. Among the 40 pairs of Drug-Drug-Interactions, 39 responses are correct for the first question, and among these 39 correct answers, 19 are conclusive while 20 are inconclusive. For the second question, 39 are correct among 40 pairs, with 17 answers conclusive and 22 answers inconclusive.

#### Administration

LLMs can serve a multifaceted role in the realm of healthcare and administrative tasks. Specifically, ChatGPT proves instrumental in streamlining administrative processes by generating texts, thereby alleviating the associated workload [15]. Moreover, it can be used to track patients’ health status, particularly those with chronic diseases [70]. Through the analysis of social media slang, GPT-3 aided in developing a drug abuse lexicon that was aimed at enhancing the monitoring of drug abuse trends [71]. Notably, an LLM-based Chatbot, called CLOVA CareCall built by NAVER AI [2], was applied as a health data-collecting tool in South Korea. Designed for emotionally supporting socially isolated individuals, CareCall conducted periodic conversations, generating health reports with metrics like meals, sleep, and emergencies. Implemented in 20 cities by May 2022, it targeted solitary adults, notably those with lower incomes, and was proven effective in reducing loneliness. Social workers used the generated reports and call recordings to monitor users’ health, resulting in positive feedback and a streamlined workload for public health workers.

### Concerns

Most of the reviewed research papers pointed out technical and ethical concerns that people harbor with respect to the application of LLMs in healthcare from several perspectives. This can generally be categorized into four groups: 1) reliability, 2) bias, 3) privacy, and 4) public acceptance.

#### Reliability

The reliability of LLMs is essential to their application in healthcare. It can be related to accuracy, consistency, and interpretability of LLM responses, and the quality of the training dataset. Specifically, 100% of prediction-related studies, 72% of summarization-related studies, and 93% of studies related to medical knowledge inquiries have reliability concerns (See Supplemental Table 1).

### Accuracy

Several studies highlighted that ChatGPT exhibited inaccuracies when asked to respond to certain questions [14,18,23,29,32,34,35,38,43,50,52,53,64,65,67,71,72]. For instance, ChatGPT could respond with incomplete information or exhibit an inability to distinguish between truth and falsehood [21,69]. The generative nature of the LLM algorithms will likely fabricate a fake reference to substantiate false claims [31], a process that has been referred to as “hallucinations” [59]. Additionally, such hallucinations can be communicated via persuasive prose [42], making it more likely to mislead patients. For example, Jo et al. mentioned that LLMs (specifically CareCall based on NAVER AI in this paper) may make ambitious or impractical promises to patients, which may add extra burden to therapists or cause a trust crisis [2].

### Data Quality

The unreliability of LLMs may be attributed to limitations in data collection sources [58,49]. There are concerns about the model’s limitation in medical knowledge [37] since the general-purpose nature of ChatGPT may affect its reliability in self-diagnosis [3]. Recent state-of-the-art LLMs are typically constructed on texts from the Internet rather than verified resources about health and medicine [1].

Of greater concern is data availability. Healthcare institutions have shared no identifiable health information with widely accessible LLMs like ChatGPT due to privacy concerns and legal compliances [7] and it is arduous to collect new data for LLM training [57]. ChatGPT, for example, was not trained on patients’ clinical data [4]. While a description of a clinical scenario without sensitive patient information can be fed into ChatGPT through prompts, it may lead to inaccurate responses [4].

Another contributing factor to inaccuracy is the outdated knowledge base used to train LLMs [21,25,30,41]. ChatGPT based on GPT3.5 was pre-trained by using data collected until 2021 and does not support Internet connection [49], making it unable to perform appropriately on questions regarding events that happened after 2021 [42].

### Consistency

Many authors expressed concerns about the inconsistency of the responses from LLMs [21,25,30], where different answers result from various prompts of the same question [17,32,39,58,59,64,67,72]. In addition, the output of ChatGPT to the same prompt may vary from user to user [17]. This is because LLMs generate responses in a probabilistic manner [1]. Therefore, nuance in the prompts to the LLM may lead to a completely different answer [17].

### Interpretability

Interpretability is another aspect regarding the reliability of the response. A study by Cadamuro et al. [60] highlights two key issues with an LLM (particularly ChatGPT) in healthcare. Firstly, the interpretation of some normal results, regarding suspected underlying diseases, was not fully correct. Second, ChatGPT struggled to interpret all the coherent laboratory tests [60], generating superficial and incorrect responses. Indeed, ChatGPT could generate overly general answers without citing original references [20,40,42].

#### Bias

It has been noted that ChatGPT has issues with disparity and bias among different populations. In other words, because certain groups of people have financial, readability, and/or accessibility barriers using LLMs, their outcomes of using LLMs will be divergent from others. For example, ChatGPT may exert some financial disparity on the users: unlike previous versions like GPT-3.5, access to GPT-4 involves a monthly fee [41]. These constraints potentially pose financial barriers, limiting widespread adoption and use of the newer, more advanced models in healthcare applications.

Moreover, the readability of an LLM’s response may further accentuate health disparity [54]. LLMs like ChatGPT include texts from scientific websites (e.g. Wikipedia) as their training data, which makes their responses sound professional and sophisticated. However, LLMs may produce biased results [6,52], making regulations to prevent bias necessary [27,53].

Furthermore, the training data can also be biased. Since recent LLMs are trained based on human-generated texts from the Internet, they also tend to provide biased answers [4]. Besides, algorithms may reinforce current health disparities and inequities [63]. Indeed, outputs from ChatGPT have been shown to be biased in terms of gender, race, and religion [4].

#### Privacy

Privacy issues are important when training or using LLMs in healthcare settings [6,7,52,70]. All AI systems including LLMs in health settings should comply with privacy regulations, including compliance with the Health Insurance Portability and Accountability Act (HIPAA), and implement robust safeguards to ensure the protection of sensitive patient information [6,7,52]. Specifically, LLMs have three privacy problems. First, the responses from LLMs may embed training examples directly, which breaches privacy if the training examples are identifiable. Second, LLMs may be susceptible to inferential disclosure. For example, a patient’s membership in a dataset or sensitive attributes may be inferred from LLMs’ responses. Third, it may not be clear whether text data is sufficiently de-identified for the anticipated recipients (which may be anyone in the world) when training LLMs. For instance, we may be able to de-identify text in a manner that sufficiently thwarts people who are not incentivized to attack the system, but we may not be addressing recipients who run machine-assisted attacks.

#### Public Acceptance

Public acceptance, the trust of the public in the application of LLMs in healthcare, has been mentioned in one study [3]. A cross-sectional survey-based study shows that 78% of a sample of 476 participants claim that they trust ChatGPT’s diagnosis, most of whom possess a degree of bachelor’s or even master’s [3]. People are inclined to trust this new technique when using ChatGPT, partially due to the convenience of obtaining information and the patients’ inclination to search for information [3].

## Discussion

This systematic review shows that LLMs have been applied to summarization, medical knowledge inquiry, prediction, and administration. At the same time, there are four major themes of concern when using these models in practice, including reliability, bias, privacy, and public acceptance. Specifically, the most popular application (30 out of 65 papers) for LLMs was for medical knowledge inquiries, with the second most popular (25) being summarization, followed by prediction (22), and then administration (9). At the same time, most of the papers expressed concerns about reliability (55), followed by bias (16), then privacy (6), and finally public acceptance (4).

### Applications

According to our systematic review, LLMs were heavily applied in summarization and medical knowledge inquiry tasks. The former is probably due to the training method of LLMs, which focuses on its capability to summarize documents and paraphrase paragraphs. The latter is due to the inclusion of general medical knowledge in the training data. Specifically, in the category of summarization, summarizing medical notes is the type of task in which LLMs were applied the most. This is probably due to the simplicity of the task and the existence of redundancy in those notes. By contrast, in the genre of medical knowledge inquiry, taking standard medical exams is the type of task in which LLMs were applied the most. This is probably due to the existence of medical questions and answers on the Internet that have been included in the training data of some LLMs such as ChatGPT.

LLMs were applied in prediction tasks as well. Specifically, in the category of prediction, diagnosis is the type of task in which LLMs were applied but with the most reliability concerns. This is probably because diagnosis is a complex process in comparison to summarization and/or the current popular LLMs (e.g., ChatGPT) used insufficient publicly available health datasets for model training. It might also be due to poorly constructed prompts without enough accurate information. Thus, LLMs are still not likely to be suitable for generating reliable answers to uncommon questions. In the category of administration, LLMs were applied equally heavily in various tasks such as appointment scheduling, information collection, and documentation.

### Concerns

For those applications of LLMs in healthcare, the two greatest concerns are reliability and bias (including disparity and inequality). These concerns might eventually drive this application away from practical implementation.

Notably, about 85% (55 of 65) of the reviewed studies emphasized concerns about the reliability of LLMs’ responses given that it may impact a patient’s health-related behavior. The concerns of reliability arose mainly from two aspects: the quality of the training data in terms of data source and data timeliness, and the models themselves. For example, GPT-3.5 was pretrained by using data collected by September 2021, and it also does not have access to private health records. Furthermore, most data that are used to train LLMs are crawled from the Internet rather than professionally validated sources. In addition, the generative nature of LLM may result in seemly professional writing but fabricating responses. However, according to Shahsavar and Choudhury [3], people are inclined to trust this new technique, due partially to the convenience of obtaining information and the patients’ inclination to search for information.

The issue of bias (or disparity) is mentioned in about 25% (16 of 65) of our included references. LLM biases come from the training stage (e.g., biased training data and biased algorithms) and the application stage (e.g., biased user base and biased outcomes). These papers discussed biases mainly from three different aspects: financial costs, readability, and accessibility. For example, Hirosawa et al. [4] pointed out that the bias encoded in human-generated texts will make LLMs generate biased output; Lee et al. [74] concerned health disparity may result from low readability made by the sophistication of LLM wording; and Johnson et al. [56] noted that LLM algorithms tend to reinforce the health disparity and to prevent LLM algorithms from exacerbating current disparity in health.

Another concern that prevents the wide application of LLMs in healthcare is privacy. When using third-party LLMs such as ChatGPT, healthcare organizations face several privacy issues. Although no privacy breach of LLMs regarding patient information has been reported, attacks for other types of private information targeting ChatGPT have been found [75]. For example, a breach led to the exposure of users’ conversations to unauthorized parties [75]. As ChatGPT interacts with patients directly, it may gather personal health information and so may breach their privacy [7]. As a result, many medical centers do not allow researchers and healthcare providers to use raw patient data as inputs to ChatGPT and other LLMs or even ban their access to these services during work [76]. Training or fine-tuning open-source LLMs requires a large number of clinical data, which may lead to violations of patients’ privacy, perhaps inadvertently [6,14,52].

### Limitations of the Reviewed Papers

The reviewed papers demonstrated two common limitations of their approaches. First of all, almost all the studies relied on human experts to rate LLMs’ responses. This is problematic because the score may be subjective and/or more likely unrepresentative. Correspondingly, future works can focus on designing a formal and fair process to evaluate LLMs’ responses from a broad range of stakeholders, including researchers, healthcare providers, patients, or any users with diverse medical and sociodemographic backgrounds. Second, some of the concerns mentioned in this review (e.g., bias) are merely researchers’ speculations of the potential risks that were included to provide directions for further work. However, the mechanisms of how the training of LLMs leads to such concerns have not been comprehensively examined through experiments. It is suggested the audience should be wary of taking these concerns for granted or as proven facts.

### Opportunities

Among all the included papers, few of them propose solutions to improve the reliability of LLMs. First, future research work should focus more on how to improve the accuracy of LLMs’ responses in the healthcare domain. More specifically, domain-specific health data are demanded for training and fine-tuning of LLMs to improve the performance of LLMs in various tasks in the healthcare domain. Therefore, data harmonization and consortia established for LLM training are potential directions that can benefit the broad research community. Qualified medical professionals can contribute to the creation of the dataset for LLM training. This, however, will be expensive in terms of time and effort [2]. Alternatively, using retrieval augmented generation (RAG) to augment LLM with external knowledge that is up to date might be a solution for scenarios where accurate in-depth professorial knowledge is required. Second, to prevent the hallucination issue, LLMs should be limited to making responses based on validated references. Blockchain technology can be used in this process to provide validation and traceability. Moreover, a holistic system, or a keep-experts-in-the-loop framework, that efficiently facilitates the expert validation process becomes important in order to improve the accuracy and safety of health LLMs. Third, clinical trials based on health outcomes such as mortality and morbidity rates should be conducted in clinical settings to validate the utility of LLM applications formally [1].

How conversational LLMs lead to bias or privacy issues in healthcare research was not thoughtfully examined with experiments in our reviewed papers. Future studies should first focus on investigating the mechanisms of how LLMs caused bias and privacy issues with stringent experiments and then developing practical solutions.

Regarding bias issues, it is suggested that systematic monitoring is necessary to ensure the impartial functioning of LLMs. However, all these sources discuss bias only with mere sentences and superficial summaries without any experimental investigation. Hence, it is worth noting that further work should also focus more on conducting experiments to understand how bias impacts the responses of LLMs in information, diagnosis, recommendation, and surveillance. More specifically, all applications of LLMs in healthcare should be tested regarding the exhibitions of bias, and the bias mitigation strategies such as data augmentation and targeted recruitment (e.g., the All of Us Research Program targets the collection of data from historically underrepresented populations [77]).

Regarding privacy issues, two technical approaches to mitigate the privacy risk while training LLMs are data anonymization [78] and synthetic data generation [79]. For deep learning models, model inversion attacks can potentially infer training data giving model weights [80]. Considering the exponentially increased open-sourced LLMs with published model weights, a sensitive patient dataset needs to be de-identified [81] or replaced with a synthetic dataset before being used to train or fine-tune an LLM. Otherwise, the patients with whom the data are associated should be informed about their participation in the training or fine-tuning process [82]. To solve the privacy issues, legal, social, and technical protection approaches need to be implemented together to ensure the privacy and security of the whole process of training and using LLMs for healthcare applications.

To raise the public acceptance level of LLMs, explainable AI should be employed to address the interpretability issues of LLMs by making the training data and model architecture transparent. More rigorous experimental studies using LLMs are encouraged in the “AI in medicine” research community to demonstrate or improve the reliability of LLM applications in healthcare. Moreover, stakeholders and decision-makers can propose new policies or regulations to manage the accountability and transparency of AI-generated content including the responses from LLMs.

There appears to be research that is beginning to address some of these raised issues. For example, Zack et al. assessed the potential of GPT-4 to perpetuate racial and gender biases in healthcare [83]. Hanna et al. assessed racial and ethical bias of ChatGPT in healthcare-related text generation tasks [84]. However, more research studies in these directions are needed to validate these findings and conduct more comprehensive and transparent assessments.

Moreover, almost all the research studies LLMs’ responses in one language. For example, 62 out of 65 study English, one focuses on Korean [2], one focuses on Chinese [45], and one focuses on Japanese [34]. Their findings cannot be extrapolated to other languages directly. Considering that many patients or people around the world or even in the US do not speak English, it is necessary to guarantee that LLMs are usable universally or equitably and conduct more research to investigate the performance of LLMs in other languages.

### Limitations of this Review

Despite notable findings, this review has several limitations. Firstly, the review used PubMed, ACM Digital Library, and IEEE Xplore as the primary sources for the papers. Other sources, such as Scopus, Web of Science, and ScienceDirect, may provide additional candidate papers regarding LLMs for Health. However, since PubMed is the main digital library for medical publications, the research findings of this review should be valuable to healthcare researchers or policymakers. Secondly, although this review intended to study the application of state-of-the-art conversational LLMs in healthcare, most of the papers included are about ChatGPT. This is because ChatGPT is still the most powerful conversational LLM. However, its closed-source nature, which is against its company name - OpenAI, may hurdle its wide application in healthcare, due primarily to the privacy concern when sharing sensitive patient information within prompts with OpenAI. Finally, only peer-reviewed papers published before September 2023 are included in our review. As a result, on the one hand, the latest LLM application developments in this area are not included in this review. Specifically, papers focused on LLMs other than ChatGPT, such as LLaMA, were very limited in our initial keyword search results, and only a few of them are included in this review. This is a problem because, while mono-modal conversational LLMs have been applied to many fields in healthcare, the multi-modal LLMs that can process medical images, such as GPT-4, Large Language and Vision Assistant (LLaVA) [85] based on LLaMA, and LLaVA-Med [73] based on LLaVA, were just released before September 2023 and are still being examined by researchers regarding their capabilities in healthcare research. As a result, no peer-reviewed research papers about applications of multi-modal LLMs in healthcare have been published before September 2023. The main challenge of the application of multi-modal LLMs in healthcare is that multi-modal LLMs are still not perfect either due to insufficient training data or due to insufficient model parameters. Specifically, with the development of computing power, reduced computing cost, and reduced data access cost, LLMs can be applied to multimedia-based diagnosis and analysis in radiology and other departments. On the other hand, the latest studies addressing the concerns are not included in this review. Although there is research that is beginning to address some of the issues raised in the systematic review [83,84], there may not have been sufficient time for all recent papers to be deposited into the repositories upon which this investigation relied yet.

### Conclusions

This review summarized applications of the state-of-the-art conversational LLMs in healthcare and the concerns that need to be resolved in the future. According to the reviewed research articles, conversational LLMs perform well in summarizing health-related texts, answering general questions in healthcare, and collecting information from patients. However, their performance is relatively less satisfying in making diagnoses and offering recommendations based on patients’ symptoms and other information. Most authors were concerned about the accuracy and consistency of the LLM responses, which should be the primary issues that researchers need to address in the near future. Nevertheless, other concerns regarding bias and privacy issues also prevent conversational LLMs from being broadly applied in the healthcare domain. However, these concerns still receive insufficient attention: few studies examine the bias and privacy issues in LLMs health-related applications with rigorous scientific experiments. Future research should focus more on conducting such research to investigate the mechanisms of how the training and application of conversational LLMs leads to such concerns, and to address these concerns that have been seen on any AI tools so that they can be safely applied in the healthcare domain.

## Data Availability

All data produced in the present work are contained in the manuscript.

## Acknowledgements

LW, ZW, and ZY conceived and designed the study. LW, ZW, CN, QS, YL, and ZY participated in paper screening and summarization. EWC, BAM, and ZY supervised the paper screening, summarization, and discussion. LW, ZW, and ZY wrote the original draft. All authors wrote the manuscript. All authors read and approved the final manuscript.

This research was funded, in part, by the following grants from the National Institutes of Health: RM1HG009034 (to EWC and BAM) and R37CA237452 (to ZY).

## Abbreviations

ChatGPT: Chat Generative Pre-trained Transformer
LLMs: large language models
NLP: natural language processing
ACM: Association for Computing Machinery
IEEE: Institute of Electrical and Electronics Engineers
PRIMSA: Preferred Reporting Items for Systematic Reviews and Meta-Analyses
LLaMA: Large Language Model Meta AI
LaMDA: Language Model for Dialogue Applications
USMLE: United States Medical Licensing Exam
OKAP: Ophthalmic Knowledge Assessment Program
AKT: Applied Knowledge Test
ESI: emergency severity index
HIPAA: Health Insurance Portability and Accountability Act
RAG: retrieval augmented generation
LLaVA: Large Language and Vision Assistant

## Footnotes

1 We use “ChatGPT-4” to represent ChatGPT Plus based on GPT-4 throughout this paper.

