## Supplemental Table 1 for "A Systematic Review of ChatGPT and Other Conversational Large Language Models in Healthcare"

**Supplemental Table 1.** A summary of applications and concerns of reviewed research papers. LLMs: large language models; DOI: digital object identifier; GPT: Generative Pre-trained Transformer; N/A: not available; AKT: Applied Knowledge Test; USMLE: United States Medical Licensing Exam; NHS: National Health Service; DR.BENCH: Diagnostic Reasoning Benchmark; EQIP: Ensuring Quality Information for Patients; VTQ: valid therapy quotient; NCCN: National Comprehensive Cancer Network; OKAP: Ophthalmic Knowledge Assessment Program; JSH: Japanese Society of Hypertension; TRS: treatment-resistant schizophrenia; LLaMA: Large Language Model Meta AI; CDS: Clinical Decision Support.

| | First Author | Application | | Concern | | LLMs | DOI |
|---|---|---|---|---|---|---|---|
| | | Category | Note | Category | Note | | |
| **1** | Akhter HM | Summarization | The authors used ChatGPT to generate case report for a patient developed with uncommon complication. | Reliability (Accuracy, Data Quality - Data Timeliness) | This paper shows that ChatGPT sometimes cites non-existent sources and is currently limited in critically discussing results and literature. | ChatGPT | 10.7759/cureus.34752 |
| **2** | Almazyad M | Summarization | This paper shows that ChatGPT can summarize conference medical recommendations. | Reliability (Accuracy), Bias (Biased training data) | This paper shows that results from ChatGPT can be biased. | ChatGPT-4 | 10.7759/cureus.38249 |
| **3** | Bhattacharyya M | Summarization | The authors investigated the authenticity and accuracy of references in ChatGPT-generated medical articles | Reliability (Accuracy) | This paper shows that ChatGPT often generates fabricated or inaccurate medical references, with a high prevalence of errors in reference elements. | ChatGPT-3.5 | 10.7759/cureus.39238 |
| **4** | Bosbach WA | Summarization | In this paper, ChatGPT drafts competent radiology reports with high appraisal given command files as input. | Reliability (Accuracy) | In this paper, ChatGPT showed limitations in its ability to deal with technical/medical terminology. | ChatGPT | 10.1067/j.cpradiol.2023.04.001 |
| **5** | Chen X | Summarization | In this paper, ChatGPT is utilized to construct a risk factor database for diseases, demonstrating its potential to extract data from literature abstracts effectively. | Reliability (Accuracy) | This paper discusses the challenges in accurately extracting risk factor information, emphasizing the need for human validation to ensure model accuracy. | ChatGPT | 10.7189/jogh.13.03037 |
| **6** | Lahat A | Summarization | This paper discusses ChatGPT's potential in formulating gastroenterology research questions. | N/A | N/A | ChatGPT | 10.1038/s41598-023-31412-2 |
| **7** | Puthenpura V | Summarization | The case report utilizes ChatGPT to integrate AI-generated text with original author writing. | Reliability (Interpretability, Accuracy) | This paper notes the potential for AI-generated text to be inaccurate, provides non-existent references in information. Dependence on AI tools | ChatGPT | 10.7759/cureus.36408 |

| | | | | | | | |
|---|---|---|---|---|---|---|---|
| | | | | | may lead to overlooking of subtle clinical signs by clinicians. | | |
| **8** | Robinson A | Summarization | ChatGPT's effectiveness in drafting operation notes for appendicectomy is evaluated in this paper, demonstrating adherence to NHS surgical documentation guidelines. | Reliability (Consistency) | This paper notes the dependency of ChatGPT's output quality on the prompt and emphasizes the need for secure integration with health records for surgical documentation. | ChatGPT | 10.7759/cureus.40546 |
| **9** | Zhou Z | Summarization | The authors explored ChatGPT's capabilities in generating medical reports from lab results with the goal of streamlining the report generation process. | Reliability (Accuracy) | This paper raises concerns about the precision and reliability of medical report generation by ChatGPT. | ChatGPT | 10.7759/cureus.37589 |
| **10** | Guirguis CA | Summarization | The authors used ChatGPT to generate case report for a patient with neurosarcoidosis. | N/A | N/A | ChatGPT | 10.7759/cureus.37368 |
| **11** | Haemmerli J | Summarization (Case Report), Prediction (Diagnosis, Treatment Recommendation) | The authors evaluated ChatGPT's performance in brain glioma adjuvant therapy decision-making. It can enhance medical case reporting and study data analysis for manuscript production. | Reliability (Accuracy, Data Quality – Data timeliness), Bias (Biased Algorithm) | This paper shows that ChatGPT is less accurate, more biased in the interpretation of medical information. It also shows that ChatGPT lacks live internet access and access to research databases. | ChatGPT | 10.1136/bmjhci-2023-100775 |
| **12** | Lyu Q | Summarization (Clinical Notes) | This paper shows ChatGPT’s ability of translating radiology reports into plain language with nice result. | Reliability (Accuracy, Consistency) | This paper shows concerns regarding ChatGPT’s moral and legal issues. For example, ChatGPT tends to oversimplify or overlook and omit key points during translation, resulting in inaccuracy. | ChatGPT, GPT-4 | 10.1186/s42492-023-00136-5 |
| **13** | Cunningham AR | Summarization, Medical Knowledge Inquiry | This paper uses ChatGPT to aid in manuscript synthesis of a patient with glioblastoma in the pineal gland exhibited over five years of survival following radiotherapy and temozolomide. | Reliability (Accuracy, Data Quality - Date timeliness, Interpretability), Bias (Readability) | This paper shows that ChatGPT cannot substitute professional medical advice and concerns about its knowledge cutoff of 2021 and its inability to access the internet. | ChatGPT | 10.7759/cureus.36590 |
| **14** | Golan R | Summarization, Medical | The authors used ChatGPT to evaluate the quality and readability of online medical text regarding | Reliability (Accuracy), | This paper shows that ChatGPT, in its current state, is less effective than human reviewers and Readable.com. | ChatGPT | 10.7759/cureus.42214 |

| | | | | | | | |
|---|---|---|---|---|---|---|---|
| | | Knowledge Inquiry | shockwave therapy for erectile dysfunction | Bias (Readability) | | | |
| **15** | Hamed E | Summarization, Medical Knowledge Inquiry | This study integrates ChatGPT-4 with “Link Reader” for automating medical text synthesis, improving AI models' traceability and retrieval accuracy. | N/A | N/A | ChatGPT-4 | 10.7759/cureus.41916 |
| **16** | Grewal H | Summarization, Medical Knowledge Inquiry, Prediction (Treatment Recommendation) | This paper highlights ChatGPT's applications in radiology, including report generation, template creation, patient communication, clinical decision-making enhancement, research title suggestion, scholarly article heading creation, and formatting and referencing for research papers. | Reliability (Data Quality - Data source, Data Timeliness), Privacy | This paper shows that ChatGPT is limited by biases in its training data, may produce inaccuracies and integrating it with EHRs risks. It may further be hampered by outdated training data. | ChatGPT (GPT-4) | 10.7759/cureus.40135 |
| **17** | Kumari KS | Summarization, Medical Knowledge Inquiry, Prediction (Treatment Recommendation) | This paper incorporates information from ChatGPT in the treatment planning and case report writing | Reliability (Accuracy), Public Acceptance | This paper shows that ChatGPT provides more general information. | ChatGPT | 10.7759/cureus.38683 |
| **18** | Cadamuro J | Summarization, Prediction (Diagnosis) | This paper evaluates ChatGPT with lab reports for relevance, correctness, helpfulness, and safety, suggesting that it can interpret individual tests but not an overall diagnostic picture. | Reliability (Accuracy, Interpretability) | In this paper, ChatGPT incorrectly interpreted normal results for suspected diseases and struggled to synthesize all related lab test findings coherently. | ChatGPT | 10.1515/cclm-2023-0355 |
| **19** | Jiang LY | Summarization, Prediction (Treatment Recommendation) | The authors developed NYUTron, a large language model trained on unstructured clinical notes for clinical predictive tasks. | Reliability (Accuracy, Consistency), Bias (Accessibility) | This paper shows that it is hard to ensure the accuracy and reliability of predictions in a clinical setting. The model lacks Generalizability. | NYUTron | 10.1038/s41586-023-06160-y |
| **20** | Sharma B | Summarization, Prediction (Diagnosis) | The authors developed DR.BENCH, a generative AI framework for clinical diagnostic reasoning tasks. They showed that | N/A | N/A | DR.BENCH | 10.18653/v1/2023.clinicalnlp-1.10 |

| | | | | | | | |
|---|---|---|---|---|---|---|---|
| | | | a multi-task, clinically trained language model significantly outperforms general domain models. | | | | |
| **21** | Liu J | Summarization. Prediction (Diagnosis, Treatment Recommendation), Medical Knowledge Inquiry, Administration (Documentation) | The authors explored ChatGPT's roles in clinical practice, focusing on clinical decision support, question-answering, and medical documentation. | Reliability (Accuracy), Privacy | The authors pointed out that potential negatives like privacy, ethics, bias, and discrimination, compounded by outdated training data, can't be overlooked. | ChatGPT | 10.2196/48568 |
| **22** | Hamed E | Summarization (Clinical Notes), Administration (Documentation) | This paper demonstrates ChatGPT's ability to adapt clinical guidelines. | N/A | N/A | ChatGPT | 10.7759/cureus.38784 |
| **23** | Kim HY | Summarization, Administration (Documentation) | This paper introduces a case study that shows that ChatGPT can help with medical documentation. | N/A | N/A | ChatGPT | 10.7759/cureus.36830 |
| **24** | Macdonald C | Summarization, Administration (Documentation) | This paper demonstrates that ChatGPT can write a paper giving a dataset. | Reliability (Accuracy) | The authors pointed out that ChatGPT can produce incorrect references and pass plagiarism detectors with 100% score. | ChatGPT | 10.7189/jogh.13.01003 |
| **25** | Cascella M | Summarization, Administration (Documentation, Information collection) | This paper shows that ChatGPT can summarize information, list possible research topics, and write clinical notes. | N/A | N/A | ChatGPT | 10.1007/s10916-023-01925-4 |
| **26** | Ali MJ | Medical Knowledge Inquiry | This paper shows that ChatGPT can provide information about lacrimal drainage disorders. | Reliability (Accuracy) | This paper shows that the information from ChatGPT is not all correct. | ChatGPT | 10.1097/IOP.0000000000002418 |

| | | | | | | | |
|---|---|---|---|---|---|---|---|
| **27** | Antaki F | Medical Knowledge Inquiry | This paper evaluates ChatGPT's proficiency in answering ophthalmic questions, showing promising results in a simulated OKAP exam. | Reliability (Accuracy, Data Quality - Data Source) | This paper shows that ChatGPT's accuracy depends on concordance and insight, with inaccuracies often due to insufficient training. | ChatGPT, ChatGPT Plus | 10.1016/j.xops.2023.100324 |
| **28** | Bird JJ | Medical Knowledge Inquiry | The authors optimized a chatbot that can answer questions regarding mental health with a high accuracy. | Reliability (Data Quality – Data source) | The authors pointed out that the available data is limited, and it takes a lot of efforts to collect data. | An unnamed chatbot | 10.1145/3594806.3596520 |
| **29** | Hoch CC | Medical Knowledge Inquiry | This paper shows that ChatGPT displays a high quiz skills and accuracy in examination. | Reliability (Accuracy) | This paper shows that ChatGPT can give false answers to a substantial proportion of questions in specific otolaryngology subdomains. | ChatGPT | 10.1007/s00405-023-08051-4 |
| **30** | Holmes J | Medical Knowledge Inquiry | In this paper, large language models, including ChatGPT, are evaluated on radiation oncology physics, with GPT-4 exhibiting superior performance and reasoning abilities. | Reliability (Consistency, Accuracy) | This paper highlights ChatGPT's consistency in answering radiation oncology physics questions yet underscores the superior performance of a team of medical physicists. | ChatGPT (GPT-3.5), ChatGPT (GPT-4), Bard (LaMDA), and BLOOMZ | 10.3389/fonc.2023.1219326 |
| **31** | Hristidis V | Medical Knowledge Inquiry | In this paper, ChatGPT and Google are compared for dementia-related queries, assessing the quality and reliability of their responses. | Bias (Readability) | This paper comments on the relevance and readability of responses from ChatGPT and Google for dementia-related queries, noting challenges in both platforms. | ChatGPT | 10.2196/48966 |
| **32** | Johnson SB | Medical Knowledge Inquiry | The authors assessed ChatGPT's ability of answering cancer information-related questions, indicating that ChatGPT provides accurate information about common cancer myths and misconceptions. | Reliability (Accuracy, Data Quality - Data Timeliness), Bias (Biased algorithm) | The authors advocated that future evaluation of AI platforms needs infrastructure to monitor for bias and health disparities, considering user trust and credibility in AI responses. | ChatGPT | 10.1093/jncics/pkad015 |
| **33** | Kung TH | Medical Knowledge Inquiry | This paper investigates ChatGPT's capability to surpass USMLE's passing threshold, showing its increasing accuracy and potential in medical education. | Reliability (Accuracy, Consistency) | This paper shows that AI's performance in medical examinations limited to human perception. | ChatGPT | 10.1371/journal.pdig.0000198 |
| **34** | Kusunose K | Medical Knowledge Inquiry | In this paper, ChatGPT provided accurate responses to CQs related to the JSH 2019 guidelines for the management of hypertension. | Reliability (Accuracy) | In this paper, ChatGPT did not provide accurate responses to some questions. | ChatGPT | 10.1253/circj.CJ-23-0308 |

| | | | | | | | |
|---|---|---|---|---|---|---|---|
| **35** | Lahat A | Medical Knowledge Inquiry | Evaluating ChatGPT's answers to gastrointestinal health questions, this study indicates its capacity to provide accurate information in certain areas. | Reliability (Accuracy) | This paper highlights the varying quality of ChatGPT's information and emphasizes the need for further development to enhance its utility for patients. | ChatGPT | 10.3390/diagnostics13111950 |
| **36** | Li Y | Medical Knowledge Inquiry | The authors refined LLaMA using 100,000 patient-doctor dialogues to provide medical advice. | Reliability (Accuracy) | The paper shows that the accuracy of LLMs such as ChatGPT could be significantly improved if they could generate or assess responses based on a reliable knowledge database with experiments. | LLaMA | 10.7759/cureus.40895 |
| **37** | Moshirfar M | Medical Knowledge Inquiry | GPT-4 outperforms GPT-3.5 and human experts in answering ophthalmology questions, with significant variations across different difficulty levels. | Reliability (Accuracy, Data Quality – Data timeliness), Bias (Financial costs, Accessibility) | The drawbacks of using CPT-4 include paying a monthly fee and having a knowledge cutoff of September 2021. | ChatGPT (GPT-3.5), ChatGPT (GPT-4) | 10.7759/cureus.40822 |
| **38** | Nov O | Medical Knowledge Inquiry | This paper assesses ChatGPT's answers to patient questions with healthcare providers, indicating its effectiveness in generating patient responses. | Reliability (Accuracy, Data Quality - Data source), Public Acceptance, Bias (Biased training data) | This paper shows that: ChatGPT and similar LLMs face issues like biased or incorrect responses, with automation bias and liability concerns requiring vigilant chatbot response curation. | ChatGPT | 10.2196/46939 |
| **39** | Sallam M | Medical Knowledge Inquiry | This paper shows that ChatGPT can challenge misinformation such as COVID-19 vaccine conspiracies. | Reliability (Accuracy), Bias (Biased training data) | The authors pointed out that ChatGPT only has limited knowledge by 2021 so that it is possible that it can produce biased and unreliable results. | ChatGPT | 10.7759/cureus.35029 |
| **40** | Sinha RK | Medical Knowledge Inquiry | This paper shows high accuracy of ChatGPT to solve higher-order reasoning questions in pathology. | Reliability (Accuracy, Data Quality - Data timeliness) | The authors pointed out that ChatGPT has limitations in that they have information on 2021 and future AI systems must be carefully designed, developed, and validated to ensure they provide accurate information. | ChatGPT | 10.7759/cureus.35237 |

| | | | | | | | |
|---|---|---|---|---|---|---|---|
| **41** | Thirunavukarasu AJ | Medical Knowledge Inquiry | This paper assesses ChatGPT's primary care application, showing promise but necessitating further development as indicated by its AKT performance. | Reliability (Accuracy), Public Acceptance | This paper acknowledges the potential and current limitations of ChatGPT in primary care, indicating a need for further development to reach the expertise level of qualified physicians. | ChatGPT | 10.2196/46599 |
| **42** | Van Bulck L | Medical Knowledge Inquiry | In this paper, 17 of 20 experts consider ChatGPT provides answers of a higher or equal value compared with Google search. | Reliability (Consistency) | This paper shows that ChatGPT is sensitive to nuance in the prompts. It uses outdated training data and is less transparent with its sources. | ChatGPT | 10.1093/eurjcn/zvad038 |
| **43** | Wagner MW | Medical Knowledge Inquiry | The accuracy of ChatGPT-3 in retrieving clinical radiological information is tested in this paper, cross-checking its responses with peer-reviewed references. | Reliability (Accuracy) | This paper expresses concerns about the accuracy and authenticity of ChatGPT-3's radiological information and references. | ChatGPT-3 | 10.1177/08465371231171125 |
| **44** | Walker HL | Medical Knowledge Inquiry | This study evaluates the reliability of medical information from ChatGPT-4 using the EQIP tool and comparison with clinical guidelines for five hepato-pancreatico-biliary conditions. | Reliability (Accuracy, Consistency, Interpretability), Bias (Readability) | This paper shows that ChatGPT-4 has no support for references, complicated answer, and accuracy issues. | ChatGPT-4 | 10.2196/47479 |
| **45** | Yeo YH | Medical Knowledge Inquiry | The performance of ChatGPT in responding to questions about cirrhosis and hepatocellular carcinoma is assessed in this paper, showing extensive knowledge in these areas. | Reliability (Data Quality) | This paper expresses concerns about ChatGPT's limitations in providing comprehensive and region-specific knowledge, particularly in managing cirrhosis and hepatocellular carcinoma. | ChatGPT | 10.3350/cmh.2023.0089 |
| **46** | Zhu Z | Medical Knowledge Inquiry | This paper shows that ChatGPT is able to pass the Chinese Medical Licensing Examination’s Clinical Knowledge Section. | Reliability (Accuracy), Privacy | The authors pointed out the importance of privacy, accuracy and reliability for an AI system. | ChatGPT | 10.1177/20552076231184091 |
| **47** | Altamimi I | Medical Knowledge Inquiry, Prediction (Treatment Recommendation) | In this study, ChatGPT is assessed for providing advice on venomous snakebites, offering accurate management information in simulated consultations. | Reliability (Accuracy) | This paper discusses reliability concerns of ChatGPT in providing snakebite management advice, stressing the need for updated knowledge and personalized information. | ChatGPT | 10.7759/cureus.40351 |

| | | | | | | | |
|---|---|---|---|---|---|---|---|
| **48** | Goktas P | Medical Knowledge Inquiry, Prediction (Diagnosis) | The authors gave examples of using "ChatGPT 4.0" in the field of allergy and immunology. | Reliability (Accuracy), Bias, Privacy | The authors pointed out the importance of privacy, and reliability. | ChatGPT 4.0 | 10.1016/j.jaip.2023.05.042 |
| **49** | Chervenak J | Medical Knowledge Inquiry, Administration (Information Collection) | This paper evaluates ChatGPT's performance on fertility-related clinical queries. | Reliability (Accuracy) | This paper shows that: 1) ChatGPT provides an illusion of reliability in its persuasive prose. 2) Different patient populations may interact with ChatGPT in different ways. 3) ChatGPT is not able to reliably cite sources. | ChatGPT | 10.1016/j.fertnstert.2023.05.151 |
| **50** | Agbavor F | Prediction (Diagnosis) | This research shows GPT-3-based text embeddings can differentiate Alzheimer's patients from healthy controls through speech data, suggesting early diagnostic potential. | Reliability (Accuracy) | This paper acknowledges the limited research on using large language models for early dementia diagnosis, specifically the potential of GPT-3. | GPT-3 | 10.1371/journal.pdig.0000168 |
| **51** | Hirosawa T | Prediction (Diagnosis) | This paper shows that ChatGPT-3 can generate diagnosis list for common chief complaints with a high accuracy. | Reliability (Data Quality - Data source, Interpretability) Bias (Biased training data) | The papers shows that it is unclear about the hyperparameters and training algorithms of the ChatGPT, thus it lacks transparency or interpretability. Also, ChatGPT may produce misleading and biased results. Last, ChatGPT lacks recent knowledge. | ChatGPT-3 | 10.3390/ijerph20043378 |
| **52** | Huang H | Prediction (Diagnosis) | This study shows that ChatGPT can be used in dental diagnosis. | Reliability (Accuracy), Bias (Accessibility), Privacy, | This study shows that ChatGPT may violate patient's privacy and ChatGPT cannot truly understand data and may produce biased results. | ChatGPT | 10.1038/s41368-023-00239-y |
| **53** | Karkera N | Prediction (Diagnosis) | The authors assessed various pre-trained large language models for extracting microbe-disease relationships from biomedical texts in zero-shot/few-shot contexts. | Reliability (Consistency), Bias (Algorithm bias) | This paper shows that varying outputs for identical prompts raise concerns about the model's response reliability. | GPT-3, BioGPT, BioMedLM, BioMegatron, PubMedBERT, BioClinicalBERT, and BioLinkBERT | 10.1186/s12859-023-05411-z |

| | | | | | | | |
|---|---|---|---|---|---|---|---|
| **54** | Sarbay İ | Prediction (Diagnosis) | ChatGPT's performance in emergency triage prediction is assessed in this paper, comparing its predictions with expert categories and scoring its sensitivity and specificity. | Reliability (Accuracy) | This paper notes discrepancies and inconsistencies in some cases. | ChatGPT | 10.4103/tjem.tjem_79_23 |
| **55** | Shahsavar Y | Prediction (Diagnosis) | This paper examines factors that influence users' intentions to use ChatGPT for self-diagnosis and health-related purposes, revealing a high willingness to adopt the technology. | Reliability (Data Quality), Public Acceptance | The papers notes that ChatGPT is not specifically designed for health care purposes, which may affect its suitability for self-diagnosis. | ChatGPT | 10.2196/47564 |
| **56** | Galido PV | Prediction (Diagnosis, Treatment Recommendation) | This paper shows that ChatGPT can identify patient as having TRS accurately and make treatment suggestions and identify drug side effects in the treatment recommendation. | Reliability (Consistency, Data Quality - Data source) | This paper shows that ChatGPT can be combined with commercial applications while generating answers. However, its output is influenced by incorrect input, and it lacks clinical context and the ability to request edits to input errors. | ChatGPT | 10.7759/cureus.38166 |
| **57** | Sorin V | Prediction (Diagnosis, Treatment Recommendation) | In this paper, ChatGPT is evaluated as a decision support tool for breast tumor board, showing promise in recommending management aligned with tumor board decisions. | Reliability (Accuracy, Consistency) | This paper reflects on the alignment of ChatGPT with tumor board recommendations, showing potential as a decision support tool with a 70% concordance rate. | ChatGPT | 10.1038/s41523-023-00557-8 |
| **58** | Juhi A | Prediction (Drug Synergy) | In this paper, ChatGPT is tested on predicting and explaining drug-drug interactions, aiming to enhance patient safety by providing accurate drug compatibility information. | Reliability (Accuracy) | This paper shows that ChatGPT sometimes provides incomplete information. | ChatGPT | 10.7759/cureus.36272 |
| **59** | Haver HL | Prediction (Treatment Recommendation) | In this paper, ChatGPT demonstrates a high accuracy in providing recommendation and prevention of breast cancer. | Reliability (Consistency) | This paper shows that ChatGPT is sensitive to nuance in the prompts. ChatGPT is a research “chatbot” not specially designed for medical use. | ChatGPT | 10.1148/radiol.230424 |

| | | | | | | | |
|---|---|---|---|---|---|---|---|
| **60** | Liu S | Prediction (Treatment Recommendation) | The authors compared ChatGPT-generated clinical support alerts with human-made suggestions, highlighting its potential for unique, understandable, and relevant contributions. | Reliability (Consistency, Accuracy, Data Quality – Data timeliness) | The authors showed that ChatGPT's responses vary with prompt changes, highlighting its sensitivity to different input sentences. | ChatGPT | 10.1093/jamia/ocad072 |
| **61** | Kao HJ | Prediction (Treatment Recommendation, Diagnosis) | The authors evaluated ChatGPT as a CDS tool in pediatrics, suggesting its capability to improve clinical workflow and assist in responsible decision-making. | Reliability (Accuracy) | The authors argued that AI technologies such as ChatGPT are not yet advanced enough to replace doctors in complex diagnoses or treatment planning. | ChatGPT | 10.1097/MD.0000000000034068 |
| **62** | Schulte B | Prediction (Treatment Recommendation, Diagnosis) | ChatGPT was used to identify guideline-based treatments for advanced solid tumors with a VTQ of 0.77 when being compared to NCCN guidelines. | Reliability (Accuracy, Consistency) | The authors showed that ChatGPT's accuracy and consistency are not certain. | ChatGPT | 10.7759/cureus.37938 |
| **63** | Carpenter KA | Administration (Information Collection) | In this paper, GPT-3 is used to generate a drug abuse lexicon from social media slang, aiming to improve pharmacovigilance and monitoring of drug abuse trends. | Reliability (Accuracy), Bias (Readability) | This paper acknowledges challenges in generating a reliable lexicon for drug abuse synonyms due to the variability of social media language. | GPT-3 | 10.3390/biom13020387 |
| **64** | Jo E | Administration (Information Collection) | The authors built CareCall, an AI tool built on HyperCLOVA, which aims at monitoring health conditions of socially isolated groups. | Reliability (Data Quality – Data source) | The authors pointed out that firsthand data is hard to gather since collecting personal health data may give rise to privacy issues. | CareCall | 10.1145/3544548.3581503 |
| **65** | Montagna S | Administration (Information Collection) | The authors utilized GPT-3 to develop an LLM-based chatbot to support management of patients' health data related to chronic diseases. | Reliability (Data Quality – Data source), Privacy | The paper show that LLMs lack of medical expertise and may be influenced by any bias in the data they were trained on. How to protect patient's privacy is another issue the authors pointed out in this paper. | GPT-3 | 10.1145/3582515.3609536 |
